# Trends and the Prevalence of Metabolic Syndrome and its Associated Factors among Nepalese Adults in Ecological regions of Nepal-A Nationwide Cross - Sectional Survey

**DOI:** 10.1101/2024.11.14.24317337

**Authors:** Bhawana Bhandari, Bihumgum Bista, Suna Laxmi Karmacharya, Anil Babu Ojha, Datin Mariani Ahmad Nizaruddin, Suresh Mehata

## Abstract

Globally, there is an increase in the Prevalence of Metabolic Syndrome. The rise in prevalence of Metabolic Syndrome suggests burden of Cardiovascular related morbidity and mortality which is also prevalent in Nepal. Metabolic Syndrome confers a 5-fold increase in the risk of Type 2 Diabetes Mellitus (T2DM) and 2-fold the risk of developing Cardiovascular Disease (CVD) over the next 5 to 10 years. Routine surveillance and registry system in Nepal related to Metabolic Syndrome are bare in Nepal which is imperative to determine the prevalence of MetS and its associated factors. This study used multistage cluster sampling considering the three ecological zones as strata. 259 Primary Sampling Units were selected randomly and a total of 5593 population were surveyed where 5051 population met the criteria for determining the Prevalence of MetS. A STEPwise Surveillance was conducted using WHO Steps survey questionnaire 3.2 version by interview technique, physical and biochemical measurements. The result concluded that there was a decrease in trend of Prevalence of Metabolic Syndrome from 14.1% to 6.69%. from 2013 to 2019.The overall Prevalence of MetS was found to be 6.69%, nearly double in women than in men (8.62% and 4.57% respectively). Waist circumference was found to be the most prevalent component of Metabolic Syndrome among women. Age, Sex, Education, Marital status, Wealth quintile and Ecological region were significantly associated with MetS and there was no significant association of MetS with behavioral factors.

## Introduction

Globally, present era is recognized for the burden for (NCDs) Non Communicable Disease [1]. Out of 51 million deaths, 41 million people die each year due to Non Communicable Disease equivalent to 71% of total deaths worldwide. Annually 17.9 million deaths occur due to cardiovascular disease which is the top most cause of death related to NCD [2]. In context to Nepal, the burden for NCDs is also high, where 66% of total deaths related to NCDs, 16% of total mortality is related with cardiovascular disease like Myocardial Infarction (MI). In 2017, 59% of total (9,015,320) DALYs are due to NCDs [3].Behavioral risk factors including smoking, harmful use of alcohol, unhealthy diet and physical inactivity, low fruits and vegetable intakes along with Metabolic/biological risk factors such as raised blood pressure (BP), blood glucose and cholesterol level, overweight and obesity have been evidenced as the rich underlying causes of NCDs [4]. The Global Burden of Disease 2017 study reported that disease burden in Nepal could be reduced by addressing these risk factors [3].

Metabolic syndrome (MS) is defined as a cluster of metabolic risk factors including central obesity, dysglycemia, reduced high-density lipoprotein cholesterol (HDL-c), elevated triglycerides (TG) and hypertension [5]. MetS are at 2-to 4-fold increased risk of stroke, a 3-to 4-fold increased risk of Myocardial Infarction (MI), and 2-fold the risk of dying from such an event compared with those without Metabolic Syndrome [6]. The massive (and increasing) disease burden associated with cardiovascular disorders justifies public health measures such as screening, which may assist with the identification and treatment of individuals who have a high risk of developing these disorders [7]. Lifestyles, behaviors and genetic factors may contribute to the prevalence of MS [8]. Interventions that address obesity and reduce waist circumference and an appropriate diet may reduce the incidence of the Metabolic Syndrome in adults[9].Screening is essential method for prior detection to identify the individual at risk to prevent from mortality and morbidity associated with Metabolic Syndrome[10].

Nepal lacks routine surveillance and registry system related to Metabolic Syndrome where baseline data are not enough to address in policy level which addresses its importance to analyze and determine the burden of MetS [11]. MetS remains unfocussed due lack of evidence at population level. At present, evidence based study of Mets representing the national level population comes to focused studies to prevent, minimize and manage the global burden of cardiovascular disease associated morbidity and mortality. Hence, the study aims for determining the trend and prevalence of MetS and its associated factor to reduce the cardiovascular burden addressing its importance in policy making and implementation and also recommendation for program like WHO PEN module in effective way to manage the Non communicable disease burden.

## Materials and Methods

**Study Site:** The study was conducted in three ecological regions of Nepal (Himalayan, Hilly and Terai region). (Fig 1)

**Fig 1:**
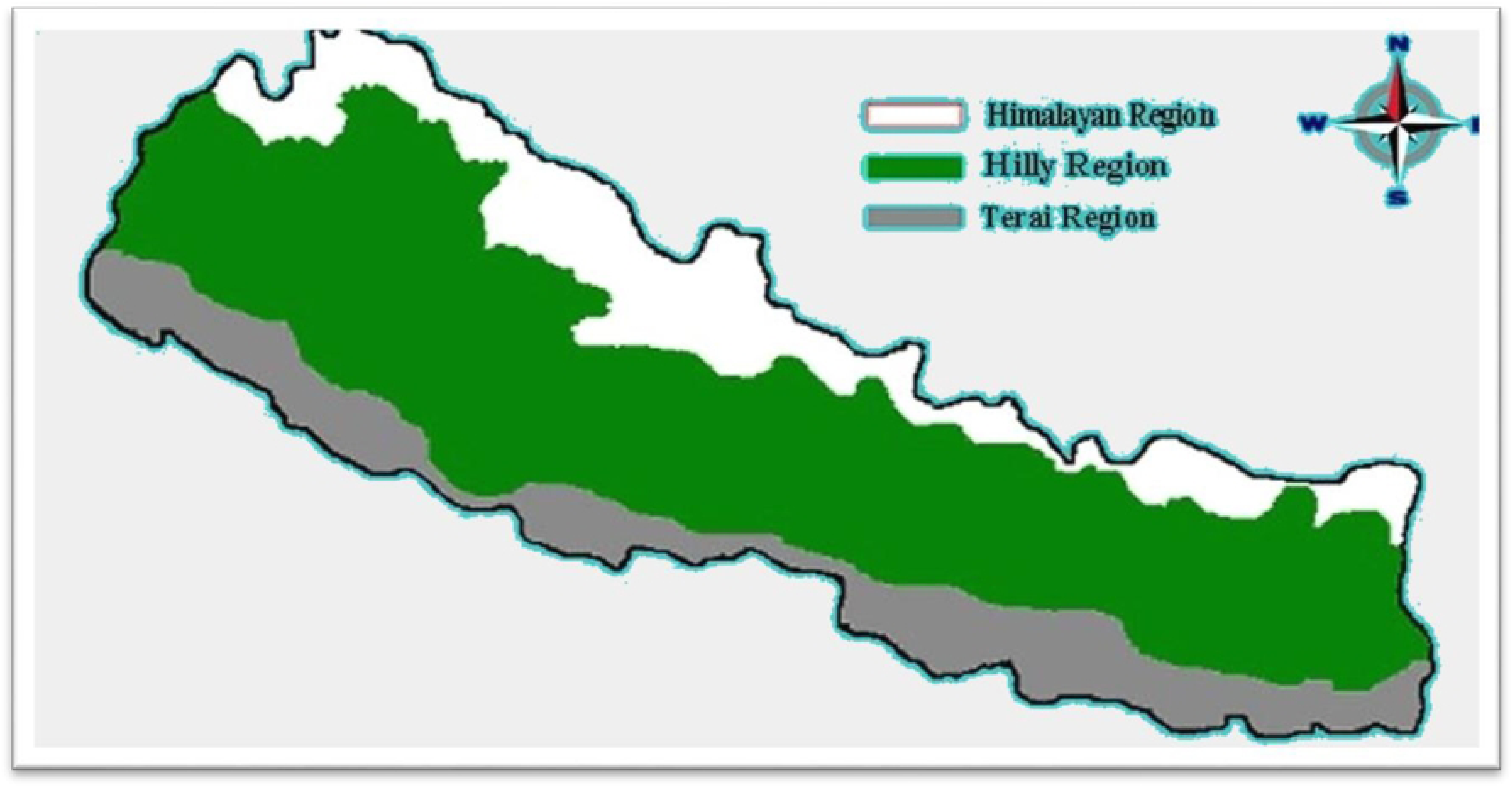

### Study design, Sampling and Data collection

This is a secondary analysis carried out using Non-Communicable Diseases risk factors Survey 2019 data. The detail methodology and report of the survey has been presented elsewhere [12]. In brief, the survey was a cross sectional study, carried out from February to May 2019 with aim to assess the risk factors of NCDs at entire population level in Nepal.

A multistage cluster sampling method was used to select 6,475 eligible participants across all 7 provinces from three ecological regions of Nepal. A total of 259 wards were selected as the primary sampling units (PSU) at the first stage, 37 PSUs from each province were selected in order to obtain provincial estimates. Secondly, household listing was carried out in 259 PSUs and list of the household was prepared.25 household per PSUs were selected using systematic random sampling. From each of the selected households, one adult member of age 15–69 years was randomly selected using KISH method through personalized computer entry system via android tablet. A total of 5593 individual participated in the survey. However, 5051 individual met the criteria for determining MetS. The response rate for STEPI, STEPII, STEPIII Survey was 86.7% (5593), 86.5%(5582) and 82.6%. (5350).Further details on the sampling process can be found elsewhere [12, 13].

We conducted face to face interviews using standardized questions from the WHO STEPS Survey (version 3.2) [14]–an update on the 2013 STEPs survey. STEP I survey collected information related to behavioral characteristics (tobacco use, alcohol use, physical activity, fruits and vegetables intake), STEP II Survey involves physical characteristics (height, weight and BP) and STEP III collected biochemical measures (Blood sugar, sodium level measurement in urine). Measurement of height (measured with portable standard stature tape SECA, Germany), Weight (measured using SECA weighing machine, Germany), BP (measured using OMRON BP monitor), blood sugar (measured using Cardiocheck PA) and blood cholesterol (measured using Cardiocheck PA) were made as per the WHO standards and STEPS manual. Household wealth was assessed on the basis of selected household characteristics (e.g. type of roof, access to electricity), means of transportation used and possession of selected consumer goods) which was used as indicator of economic status. Households were given scores based on the number and kind of consumer goods they own. The scores are derived using Principal Component Analysis (PCA). Details of the measurement process has been described elsewhere [12,15].

Analysis was performed with STATA version 15.1 using survey (*svy*) set command, defining clusters and sampling weight information. All estimates were weighted by sample weights in order to obtain the population estimates and are presented with 95% Confidence Intervals (CI). Prevalence estimates were calculated using Taylor series linearization. The trend in the study is the state of Prevalence of Metabolic Syndrome for increase, decrease or static. For this, the Prevalence Rate will be compared with that of 2013 STEPS survey data to 2019 STEPS survey data. Chi-square tests were used for bivariate analysis, to test associations between independent and dependent variables. Furthermore, multivariate Poisson regression was used to calculate the Adjusted Prevalence Ratio (APR) between each risk factors of Metabolic Syndrome and socio-demographic covariates (age, sex, education, marital status, province, ecological regions and place of residence) included simultaneously [16]. For clustering analysis of Metabolic Syndrome the numbers of risk factors present within each participant were summed (from 0 to 4). Bivariate analysis and multivariate analysis were analyzed against socio-demographic covariates through Poisson regression. The relationship between the number of risk factors and covariates were estimated through Adjusted Odd Ratios (AOR), with the count of risk factors designated as the dependent variable.

#### Variable definition

**The dependent variable:** Prevalence of Metabolic Syndrome.

**The independent variable**: Three major predictor variables were included:

i. **Individual characteristics**: age in years (15–29, 30–44 and 45–69), gender (male and female), marital status (never married, currently married and divorced/widowed/separated), education (no formal schooling, primary, secondary and higher level) and wealth quintile.
ii. **Community characteristics**: Ecological zone (Mountain, Hill and Terai/ plains) and place of residence (urban and rural) in this analysis.
iii. **Behavioral characteristics**: smoking tobacco, alcohol consumption, intake of fruits and vegetables and physical activity.

For this study, current smoking, harmful use of alcohol, insufficient fruit and vegetable intake, insufficient physical activity are considered as a behavioral factor. Similarly, raised Blood Pressure is categorized as a physical factor. Raised blood sugar and raised blood cholesterol together are considered as biochemical factor. The operational definitions of the outcome variables (NCD risk factor) are presented in Table 1.

**Table 1:** Showing the variables used in study.

| Variables | Parameter |
| --- | --- |
| **Prevalence:** | The occurrence of disease in a defined population in the given period of time. The total population surveyed was 5593 with 84% response rate.<br>Calculated by: $\frac{\text{Total no of people affected}}{\text{Total population at that point of time}} * 100\%$ [ 17] |
| **Waist and hip circumference** | Waist was measured using constant tension tapes (Seca, Germany) in centimeters [18].<br>Waist circumference: >102 cm (Male), >88 cm (Female) was taken into consideration for the study [19]. |
| **Blood Pressure** | Blood pressure was measured with a digital, automated blood pressure |
| **measurements:** | monitor (OMRON digital device) with medium cuff. Before taking the measurements, participants were asked to sit quietly and rest for 15 minutes with legs uncrossed[18].<br>The mean readings for sum of second and third measurements, blood pressure $\geq 130$ mmHg systolic or $\geq 85$ mmHg diastolic or Rx was considered for the analysis [19] |
| **Biochemical measurements** |  |
| **Fasting glucose** | • **Fasting glucose:** For the measurement of **fasting glucose**, participants were instructed to fast overnight for 12 hours and diabetic patients on medication were reminded to bring their medicine/insulin with them and take their medicine after providing the blood sample Fasting glucose $\geq 100$ mg/dl or Rx was taken for reference for the study [18,19] |
| **Total cholesterol** | • **Total cholesterol** level was screened for cardiovascular risk in STEPS Survey. The Normal value for Total Cholesterol level was ≥<br>5.0 mmol/l or $\geq 190$ mg/dl or Rx [18,6]<br>The NCEP recommended that HDL cholesterol be measured in persons with high total cholesterol levels or in those with borderline high levels and a high CHD risk profile [20]. Current NCEP ATPIII guidelines appear to balance optimally the sensitivity and specificity for selecting individuals based on TC levels for further complete lipoprotein analysis [21]. Elevated TC is a well-documented and established risk factor for cardiovascular disease. One large meta-analysis based on observational studies found that high levels of serum TC was associated with an increased Coronary Heart Disease mortality rate [22]. |
| **Metabolic Syndrome** | The parameters for Waist Circumference, Fasting Glucose Level and Elevated Blood Pressure for measuring Metabolic Syndrome were taken from NCEP III criteria[19].<br>Total Cholesterol parameter was taken from NCD Risk factor survey. |
|  | Thus, assessment criteria for Metabolic Syndrome include any three of four criteria.<br>• Waist circumference: >102 cm (Male), >88 cm (Female)[19]<br>• Fasting glucose ≥100 mg/dl or Rx [19].<br>• Elevated blood pressure Rx ≥130 mmHg systolic or ≥85 mmHg diastolic or Rx [19]and<br>• Dyslipidemia: Total cholesterol level>5.0 m/mol or Rx [18] |
| **Current tobacco:** | Those who use smoke and smokeless tobacco smoked in the past 30 days were considered as a current tobacco smoker [18]. |
| **Alcohol consumption:** | The participants were classified as life time abstainers, alcohol consumed in past 12 months and consumed in past 30 days [18]. |
| **Insufficient fruits and vegetable intake:** | It is the number of servings of fruit and vegetables consumed on average per day. Average serving of fruits and vegetables consumed per day 2.0 servings (0.5 servings of fruit and 1.5 servings of vegetables per day is sufficient[18]. Less than average serving was considered insufficient intake. |
| **Low Physical activity:** | Global Physical Activity Questionnaire (GPAQ) was used to assess physical activities. On an average 299.2 minutes per day is adequate whereas level of physical activity less than 150min per week is inadequate [18]. |
| **Nepalese adults:** | The adult citizen from 15-69 years of age residing in three geographical regions of Nepal. |
| **Ecological Zone:** | Ecological zone classify Nepal into 3 parts Himalayan, mountainous /hilly and terai /plains [18]. |

### Ethics statement

Ethical approval for conducting the study was taken from Nepal Health Research Council, Government of Nepal (Reference No. 3755, Regd. No. 77/2023, approved on 10^th^ May, 2023). The agreement from Nepal Health Research Council for utilizing Raw data of STEPS Survey was done on 17^th^ July 2022. Written informed consent was obtained from each participant before they enrolled in the survey. In case of minors (under 18 years old) both assent from the research participants and consent from their parents (legal guardian) was obtained, as per national ethical guidelines for health research in Nepal. We also took administrative approval from federal, provincial and local governments, as per the need. The confidentiality of all information gathered was maintained. Any waste generated during the laboratory procedures was properly disinfected using aseptic techniques before being safely disposed of. All blood and urine samples were discarded after completing biochemical measurements [18].

### Global inclusivity

Additional information regarding Metabolic Syndrome on screening measures including Total Cholesterol rather than its individual component which suggest cost effectiveness in screening of MetS was included in the study. In a previous nationally representative study using STEPS Survey 2013 data, an association of Socio-economic Status (Wealth quintile) and the Metabolic Syndrome was not performed. However, due to globalization and technological modernization, the society has been changing rapidly. We, therefore investigated the relation between wealth quintile and the Metabolic Syndrome among Nepalese adults.

## Results

**Table 2. Socio-demographic Characteristics of Respondents**

Socio-demographic characteristics of the participants are summarized in Table 2.Participants’ age ranged from 15 to 69 years. Majority (37.33%) of the participants aged from 45-69 years. More than half (64.28%) of the participants were female. Almost (84.81%) participants were married and almost half of the participants (46.59%) reside in Hilly region. 29% had poorest quintile household assets. Almost half of the participants (49.93%) had none/primary level education. Majority of the participants (87.39%) were from rural areas.

**Table 2.** Socio-demographic characteristics of respondents.

| Characteristics | Frequency (f) | Percentage (%) |
| --- | --- | --- |
| **Age (in years)** |  |  |
| 15-29 | 1,466 | 26.61 |
| 30-44 | 2,039 | 36.41 |
| 45-69 | 2,088 | 37.33 |
| **Sex** |  |  |
| Women | 3,595 | 64.28 |
| Men | 1,998 | 35.72 |
| **Level of education** |  |  |
| None/less than primary | 2792 | 49.93 |
| Primary | 1052 | 18.79 |
| Secondary | 1088 | 19.46 |
| More than secondary | 661 | 11.82 |
| **Marital status** |  |  |
| Never married | 487 | 9.7 |
| Currently Married | 4283 | 84.90 |
| Divorced/separated/widow | 280 | 5.4 |
| **Household assets (in quintile)** |  |  |
| Poorest | 1652 | 29.54 |
| Second | 1063 | 19.01 |
| Third | 948 | 16.95 |
| Fourth | 881 | 15.75 |
| Richest | 1049 | 18.76 |
| **Geographical Region** |  |  |
| Mountain | 661 | 11.8 |
| Hill | 2,606 | 46.59 |
| Terai | 2,326 | 41.58 |
| **Residence** |  |  |
| Urban | 705 | 12.61 |
| Rural | 4888 | 87.39 |

**Fig 2: Trends in the Prevalence of Metabolic Syndrome between 2013 and 2019.**

The plot in the graph below “Fig 2” shows the decrease in the trend of Prevalence of Metabolic Syndrome. The prevalence of Metabolic Syndrome is decreased to more than half which is 14.1% in 2013 to 6.69% in 2019.

**Fig 2:**
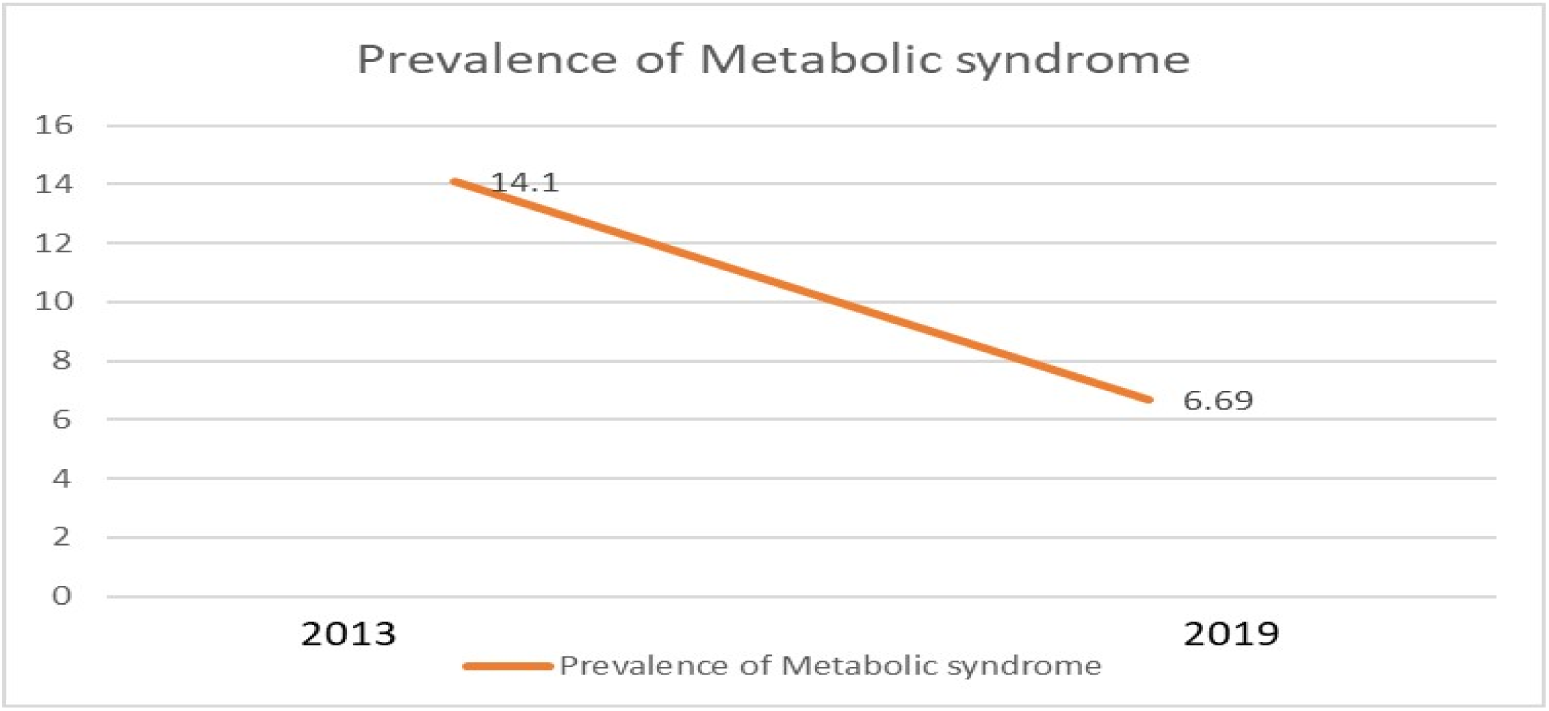
Trend in the Prevalence of Metabolic Syndrome between 2013 and 2019.

Table 3 represents the bivariate analysis of Metabolic Syndrome and Socio-demographic variables. Metabolic syndrome was higher in participants aged 45-69 years (11.95%), in women (8.62%), who has none/less than primary education (8.81%), who are divorced/separated (13.6%), participants living in Terai (7.77%) and richest (11.78%) in quintile. Metabolic syndrome was significantly associated with Age, Sex, Education, Marital status, Geographical region and Household assets in quintile p ≤ 0.05.

**Table 3.**
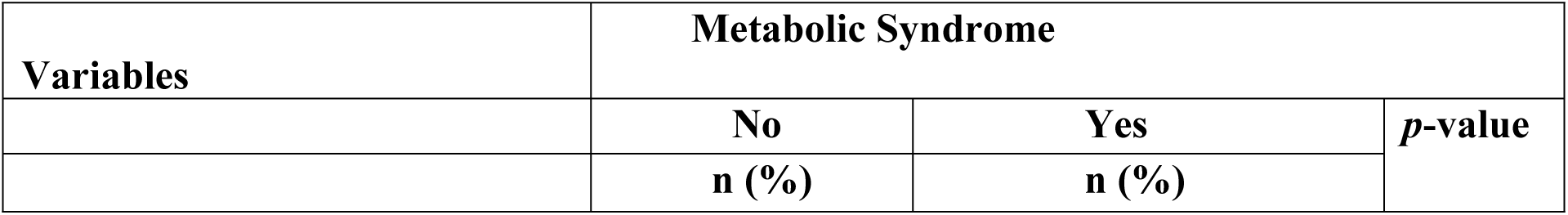
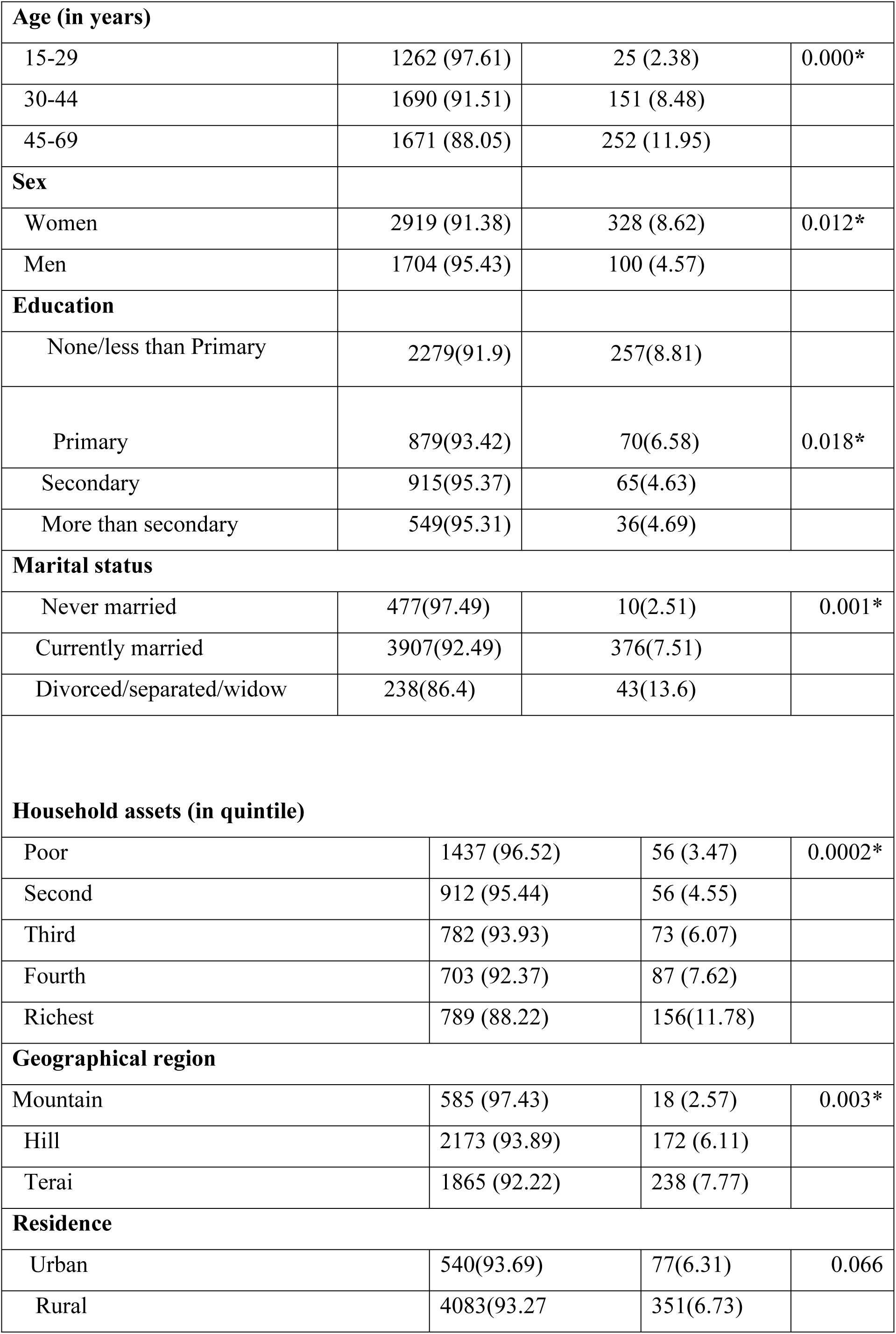
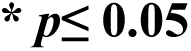
Association between Metabolic Syndrome with the socio-demographic variables.

| Variables | Metabolic Syndrome |  |  |
| --- | --- | --- | --- |
|  | No | Yes | p-value |
|  | n (%) | n (%) |  |
| **Age (in years)** |  |  |  |
| 15-29 | 1262 (97.61) | 25 (2.38) | 0.000* |
| 30-44 | 1690 (91.51) | 151 (8.48) |  |
| 45-69 | 1671 (88.05) | 252 (11.95) |  |
| **Sex** |  |  |  |
| Women | 2919 (91.38) | 328 (8.62) | 0.012* |
| Men | 1704 (95.43) | 100 (4.57) |  |
| **Education** |  |  |  |
| None/less than Primary | 2279(91.9) | 257(8.81) |  |
| Primary | 879(93.42) | 70(6.58) | 0.018* |
| Secondary | 915(95.37) | 65(4.63) |  |
| More than secondary | 549(95.31) | 36(4.69) |  |
| **Marital status** |  |  |  |
| Never married | 477(97.49) | 10(2.51) | 0.001* |
| Currently married | 3907(92.49) | 376(7.51) |  |
| Divorced/separated/widow | 238(86.4) | 43(13.6) |  |
| **Household assets (in quintile)** |  |  |  |
| Poor | 1437 (96.52) | 56 (3.47) | 0.0002* |
| Second | 912 (95.44) | 56 (4.55) |  |
| Third | 782 (93.93) | 73 (6.07) |  |
| Fourth | 703 (92.37) | 87 (7.62) |  |
| Richest | 789 (88.22) | 156(11.78) |  |
| **Geographical region** |  |  |  |
| Mountain | 585 (97.43) | 18 (2.57) | 0.003* |
| Hill | 2173 (93.89) | 172 (6.11) |  |
| Terai | 1865 (92.22) | 238 (7.77) |  |
| **Residence** |  |  |  |
| Urban | 540(93.69) | 77(6.31) | 0.066 |
| Rural | 4083(93.27) | 351(6.73) |  |

**\* $p \leq 0.05$**

Table 4 showed no any significant association of Metabolic Syndrome with Behavioural Characteristics.

**Table 4.**
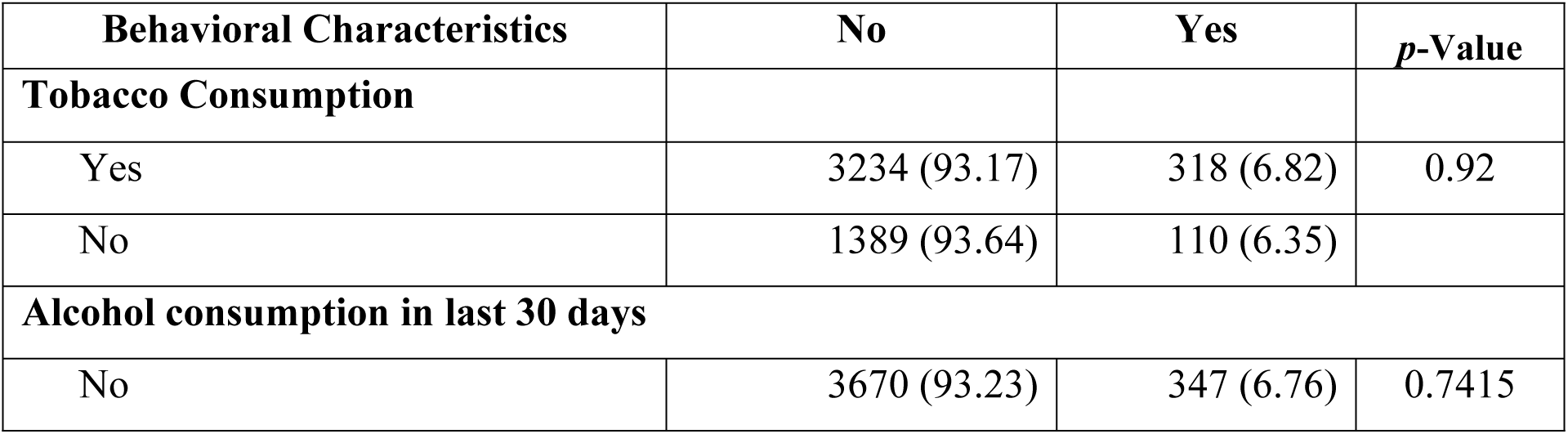
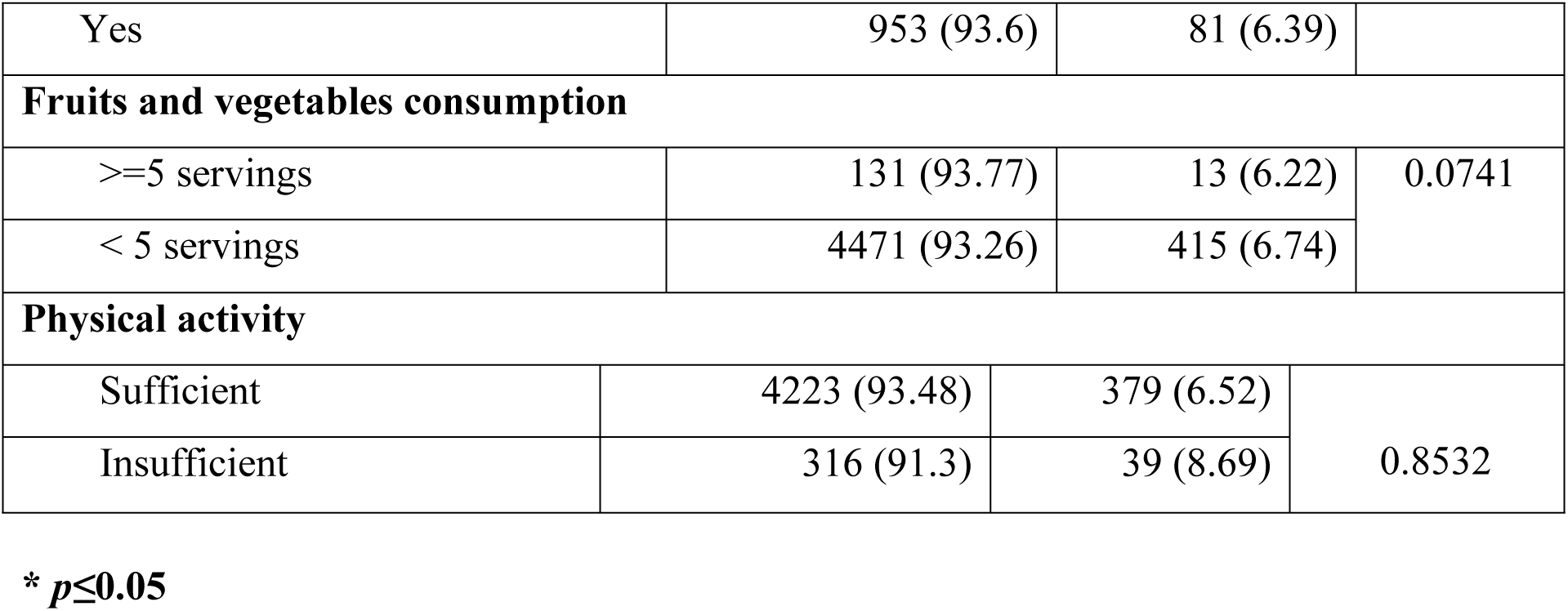
Association of Metabolic Syndrome with behavioral characteristics.

| **Behavioral Characteristics** | **No** | **Yes** | ***p*-Value** |
| --- | --- | --- | --- |
| **Tobacco Consumption** |  |  |  |
| Yes | 3234 (93.17) | 318 (6.82) | 0.92 |
| No | 1389 (93.64) | 110 (6.35) |  |
| **Alcohol consumption in last 30 days** |  |  |  |
| No | 3670 (93.23) | 347 (6.76) | 0.7415 |
| Yes | 953 (93.6) | 81 (6.39) |  |
| Fruits and vegetables consumption |  |  |  |
| >=5 servings | 131 (93.77) | 13 (6.22) | 0.0741 |
| < 5 servings | 4471 (93.26) | 415 (6.74) |  |
| Physical activity |  |  |  |
| Sufficient | 4223 (93.48) | 379 (6.52) | 0.8532 |
| Insufficient | 316 (91.3) | 39 (8.69) |  |

\* $p \leq 0.05$

**Table 5. Multivariate analysis for association of Metabolic Syndrome with socio-demographic variable**

There was a significant association between Age and Metabolic Syndrome (p<0.001). The odds of having Metabolic Syndrome are 3.68 times higher among participants aged 30-44 years (p-value: <0.001, 95% CI: 1.94,6.99) and 5.79 times higher among participants aged 45-69 years (p-value: <0.001, 95% CI: 2.77, 10.41) compared to those participants who are age of 15-29 years adjusting for sex, marital status, education, residence, geographical regions, wealth index, current use of tobacco product, alcohol consumption in last 30 days, fruits and vegetable consumption, physical activity per week. The cumulative risk for Metabolic Syndrome increased with increase in age.

There was significant association between Sex and Metabolic Syndrome (p<0.001). The odds of having Metabolic Syndrome are 2.45 times significantly higher in women (95% CI: 1.57, 3.51) adjusting for age, residence, marital status, geographical region, wealth index, current use of tobacco product, alcohol consumption in last 30 days, fruits and vegetable consumption, adequate physical activity.

The higher odds of occurrence of Metabolic Syndrome was observed among rural dwellers (AOR 1.48; 95% CI: 0.93-2.35) as compared to urban dwellers adjusting for age, residence, marital status, geographical region, wealth index, current use of tobacco product, alcohol consumption in last 30 days, fruits and vegetable consumption, adequate physical activity.

There is significant association between geographical region (Terai) (p-value: 0.02, 95% CI:1.06, 3.99) and Metabolic Syndrome. The odds of having Metabolic Syndrome is 2.06 times higher in those who live in the Terai region and 1.68 times higher in those who live in the mountainous region adjusting for age, residence, marital status, geographical region, wealth index, current use of tobacco product, alcohol consumption in last 30 days, fruits and vegetable consumption, adequate physical activity.

The odds of developing Metabolic Syndrome is 0.84 times high in those who are currently married and 1.21 times higher in those who are separated/divorced/widow, adjusting for age, residence, marital status, geographical region, wealth index, current use of tobacco product, alcohol consumption in last 30 days, fruits and vegetable consumption, adequate physical activity.

The odds of having Metabolic Syndrome decrease with increase in education level. The odds of having Metabolic Syndrome is 1.0 times more who have primary education and decrease odds with secondary education (AOR; 0.80) and higher secondary education (AOR; 0.79) compared to those who have no or less than primary education adjusting for age, residence, marital status, geographical region, wealth index, current use of tobacco product, alcohol consumption in last 30 days, fruits and vegetable consumption, adequate physical activity. The cumulative risk of metabolic syndrome decreased with increased level of education.

The odds of developing Metabolic Syndrome is 2.43 times high in those who have fourth wealth index (p-value: 0.006, 95% CI: 1.29,4.60) and 4.12 times higher in those who have highest wealth index (p-value: <0.001, 95% CI: 2.30,7.39) compared to those who have poor wealth index adjusting for age, residence, marital status, geographical region, wealth index, current use of tobacco product, alcohol consumption in last 30 days, fruits and vegetable consumption, adequate physical activity.

Age, female, education, wealth quintile Marital status and Terai residents were at increased of Metabolic Syndrome.

**Table 5.** Multivariate analysis for association between socio-demographic variables and Metabolic Syndrome.

| **Characteristics** | **Odds ratio** | **95% CI** | ***p*-value** |
| --- | --- | --- | --- |
| **Age (in years)** |  |  |  |
| 15-29 | Ref |  |  |
| 30-44 | 3.68 | 1.94,6.99*** | <0.001 |
| 45-69 | 5.79 | 2.7710.41*** | <0.001 |
| **Sex** |  |  |  |
| Men | Ref |  |  |
| Women | 2.46 | 1.58 , 3.82*** | <0.000 |
| **Residence** |  |  |  |
| Urban | Ref |  |  |
| Rural | 1.48 | 0.93,2.35 | 0.098 |
| **Marital status** |  |  |  |
| Never married | Ref |  |  |
| Currently married | 0.84 | 0.35,2.04 | 0.699 |
| Divorced/separated | 1.21 | 0.48,3.08 | 0.684 |
| **Education** |  |  |  |
| None/less than primary | Ref |  |  |
| Primary | 1 | 0.63 , 1.58 | 0.995 |
| Secondary | 0.8 | 0.48 , 1.33 | 0.379 |
| More than secondary | 0.79 | 0.38 , 1.64 | 0.518 |
| **Wealth Index** |  |  |  |
| Poorest | Ref |  |  |
| Second | 1.46 | 0.87 - 2.44 | 0.178 |
| Middle | 1.82 | 0.92 - 3.60 | 0.107 |
| Fourth | 2.66 | 1.42 - 4.99** | 0.006 |
| Richest | 4.84 | 2.65 - 8.85*** | 0.000 |
| **Geographical Region** |  |  |  |
| Mountain | Ref |  |  |
| Hills | 1.69 | 0.88, 3.25 | 0.116 |
| Terai | 2.06 | 1.08, 3.95** | 0.029 |

\* $p<0.05$ ; \*\* $p<0.01$ ; \*\*\* $p<0.001$
*Adjusted for age, sex, residence, marital status, education, residence, wealth index,* *geographical regions current use of tobacco product, alcohol consumption in last 30 days,* *fruits and vegetable consumption, adequate physical activity.*

Table 6 depicts the Prevalence of Components of Metabolic Syndrome. The overall Prevalence of Metabolic Syndrome was 6.6%. The Prevalence was significantly double (8.62%) among women as compared to men (4.2%).The most prevalent component was High Waist Circumference (63.6%) that was significantly high among women followed by Elevated Blood Pressure (29.8.%) which was significantly high among men. Total Cholesterol (10.8%) was observed in both Sex which was significantly high (13.9%) among women. Among the components observed, Raised blood sugar was least prevalent (6.3%) in both sex which was observed more (5.3%) among the women. Majority (93.4%) of respondents has less than 3 components and was significantly less among Men.

**Table 6.** Prevalence of components of Metabolic Syndrome.

| Metabolic Components | Women% | Men% | Both Sex% | *p-value* |
| --- | --- | --- | --- | --- |
| High waist hip ratio (n=5518) | 70.2 | 56.3 | 63.6 | 0.000* |
| Elevated blood pressure<br>(n=5506) | 19.7 | 29.8 | 24.75 | 0.000* |
| Elevated Total blood cholesterol<br>(n= 5342) | 13.9 | 7.7 | 10.8 | 0.000* |
| Raised blood sugar (n=5191) | 5.3 | 4.2 | 6.3 | 0.282 |
| Metabolic syndrome (n=5051) | 8.62 | 4.57 | 6.6 | 0.001* |
| 0 or < 3 components (n=5051) | 51.21 | 48.79 | 93.4 | 0.0002* |

\* $p < 0.05$

Clustering of Components of Metabolic Syndrome was observed among the respondents in Table 8. A triad of blood pressure, blood sugar and Total Cholesterol (n=219) constitutes 3.59% in both sex and was common among women (54.85%) followed by triad of waist hip ratio, blood pressure and blood sugar (n=185) that constitutes of 3% in both sex and was significantly high among women. Triad of waist hip ratio, blood pressure and total cholesterol (n=134), 1.94% was noticeable in both sex and showed significantly high among women. A triad of waist hip, blood sugar and Total Cholesterol (n=97) was least dominant i.e. 1.41% was seen in both sex and was significantly high among women. Clustering of all four components (Waist hip Circumference +Elevated Blood Pressure Raised Blood Sugar+Total Cholesterol) was seen among 1.09% of Respondents and was significantly high among Women. Hence, clustering of Components of Metabolic Syndrome components was found to be high among women.

**Table 7.** Clustering of components of Metabolic Syndrome.

| **Clustering of Metabolic Components** | **Women%** | **Men%** | ***p-value*** |
| --- | --- | --- | --- |
| Waist hip Ratio+ Blood pressure+ Total cholesterol (n=134) | 89.49 | 10.51 | 0.001* |
| Waist hip Ratio+ Elevated Blood Pressure+ Elevated Blood sugar (n=185) | 90.49 | 26.86 | 0.0067* |
| Waist hip Ratio + Elevated Blood Sugar + Total Cholesterol (n=97) | 91.49 | 14.26 | 0.000* |
| Elevated Blood Pressure+ Elevated Blood Sugar + Total Cholesterol (n=219) | 92.49 | 41.92 | 0.000* |
| Waist hip Ratio+ Elevated Blood Pressure+ Elevated Blood Sugar+ Total Cholesterol (n=66) | 93.49 | 17.03 | 0.0016* |

**\*p<0.05**

## Discussion

The present study demonstrated that the overall prevalence of Metabolic Syndrome was 6.69% which shows decrease in trend from 12.1% in 2013 to 6.69% in 2019 (Figure 2).The study is in contrast with the Prevalence of Metabolic Syndrome among South Asian countries (ATPIII 26.1% and IDF 29.8%) reported by a systematic review [23].Similarly, the study is in line with the study that showed the prevalence of the Metabolic Syndrome was lower in populations of Asian origin compared with what has previously been reported for Caucasians[24] .The decrease in trend of Metabolic Syndrome in Nepal may be related to decrease in the prevalence of Total Cholesterol in 2019. The prevalence of Raised Total Cholesterol in STEPS Survey 2019 is quite low (11%) as compared with previous rounds of the STEPS Survey in 2013 i. e. 22.7%;.[18] and also may be due to differences in measurement criteria (Table 1). Education status also play vital role in creating awareness regarding health risk.

Socio-demographic characteristics like Age, Sex, Education, Marital Status, Geographical region and Household assets in quintile have statistical significant association with Metabolic Syndrome which is consistent with the study in Nepal [25].

In the present study, Behavioural factors has no association with Metabolic Syndrome which was in line with the study conducted in USA where Lifestyle factors had no significant association with metabolic syndrome [26].

Higher Trend of Metabolic Syndrome was observed among women (8.62%) than on men (4.57%) with the most prevalent three metabolic components in women (High Waist Circumference: 70.2% of women, 5.3% of men, Elevated Total Cholesterol: 13.9% of women; 7.7% of men; Raised blood sugar 5.3% in women; 4.2% in men) (Table 7) was found to be supported by study in India [27]. Conversly, the prevalence of MetS in China was higher among men than women and also the prevalence of the four metabolic abnormalities included in the diagnosis of Metabolic Syndrome were also higher in men than in women [28]. Similar findings on the study at India, showed the pooled prevalence of MS among adult women was more 35% (95%CI: 31%-38%); than men, the pooled prevalence was 26% (95%CI: 22%-29%) [29]. These results support the existing research findings that women experience physiological changes that make them susceptible to metabolic syndrome with aging. Menopause, characterized by a decline in circulating estrogen levels, may increase cardiovascular risk through its effects on adiposity and lipid metabolism. The sex differences in Metabolic Syndrome may be related with biological and behavioral factors [30]. The factors associated with NCDs among female were due to long working hours, double work burden, and stress which keeps them busy with managing the household activities that reduces the time for physical fitness [31] .The prevalence of physical inactivity was found to be doubled in STEPS SURVEY 2019 and is found to be more in women than in men and is associated with richest wealth quintile [32],unemployment and easy access to transportation [33],lower social classes, as defined by education and occupational classification [34] and low education among females [35].

There was significant difference between Metabolic Syndrome with Age. The prevalence of Metabolic Syndrome rose progressively with age and is high enough at the forty to sixty decades of age. Prevalence of Metabolic Syndrome was found to be more with increase in age which may be due to ageing process association with increased adiposity that eventually leads to deposition of adipose fast and pathological hormonal deficiencies. The increasing pro inflammatory factors with aging are disruptive to insulin receptor signaling in adipocytes and are associated with development of diabetes and other metabolic diseases [36].The age-related adipose tissue dysfunction leads to the consequent systemic inflammation and metabolic dysfunction [37].

There was significant difference between Metabolic Syndrome with education with a lower prevalence found in most educated respondents and high prevalence was noticed in the respondents who had less than primary and the prevalence was decreased with increment in educational status. The study was related with the study done in China which showed Lower education level was associated with higher risk of MetS [38].This may be due to accessibility of awareness and concern among educated people on the health benefits and risk which help them to adopt health preventive and promotive measures [39].

Metabolic Syndrome was found to be significantly associated with Marital status. The odds of having Metabolic syndrome was high among divorced/ separated then that of ever married and currently married women which is similar with the study conducted in Korea among Korean married women where the married middle-aged group, the widowed middle-aged group tended to have a higher risk of Metabolic Syndrome, which is predicted to be related to socioeconomic factors, health behavior [40] and marital distress[41].Women in high-quality marriages are at lower risk of developing the metabolic syndrome[42].

Metabolic Syndrome is significantly associated with the fourth and richest quintile. The odds of having Metabolic Syndrome was more than double in fourth quintile and more than four times in richest quintile as compared to the poorest quintile. Various studies suggests that sedentary habits, behavior problems related to health, low prevalence of physical activity, stress and eating habit are prominent with increased in assets[18].

Metabolic Syndrome was found to be ecologically associated. There was strong association of Metabolic Syndrome with geographical region. The study was in line with the study that showed the prevalence of multi-morbidity was more in low altitude [43]. Terai region geographically lies in low altitude where culture and traditions linked with food preparation and consumption, dietary habits, adoption of behavioral risk factor, climate and stress increases the risk of NCD [44]. In contrast Urbanization, specifically rurality and high-altitude mainly ≥2,500 meters above sea level (m.a.s.l.), were factors independently associated with lower predicted Cardiovascular risk [45].

The higher Prevalence of components of Metabolic Syndrome identified in the study was high Waist Circumference which was more among the Women. In contrary, the prevalence of MetS from nationally representative population addressed the most predominant component was low HDL cholesterol among female population [25].

## CONCLUSION

It was concluded that there was a decrease in Trend of Prevalence of Metabolic Syndrome in Nepal. The prevalence of Metabolic Syndrome was considerably double in Women than in Men. Age, Sex, Education, Marital Status, wealth quintile and ecological regions are associated factors for Metabolic Syndrome. High Waist Circumference among women was the most prevalent component of Metabolic Syndrome.

## RECOMMENDATIONS

Metabolic Syndrome increases the risk of Cardiovascular disease. To decrease the mortality and morbidity associated with cardiovascular disease, screening on MetS could be helpful.

Cost effective measures should be taken to perform screening measures. Total cholesterol solely can be utilized instead of HDL and LDL that can economize the screening for MetS. The recommendations for identifying high risk individuals are based on screening measures of total cholesterol, although the recommendations for treatment are based on concentrations of LDL cholesterol. Total cholesterol was recommended as the initial screening test for three reasons: High levels of LDL cholesterol are associated with an increased risk of CHD; there is a high correlation between total cholesterol and LDL cholesterol; and testing for total cholesterol is easier and less expensive than testing for LDL. The NCEP recommended that HDL cholesterol be measured in persons with high total cholesterol levels or in those with borderline high levels and a high CHD risk profile [20]. Current health Plan could include the screening of MetS to address burden of Cardiovascular disease in WHO PEN module package to decrease the burden of cardiovascular related premature mortality and morbidity. Marital status appears to be related to cardiovascular disease in among women, it is important to provide lipid screening and educational programs to improve women’s lifestyle based on their marital status. High waist circumference among women highlights the urgency of addressing abdominal obesity in health care priority.

## Data Availability

The data are available and stored safely. The author declare that the data is original and is ready to submit the data at any time.

## Acknowledgement

The author would like to acknowledge University Grant Commission (UGC) to provide partial scholarship for conducting this study. The authors would like to offer gratitude to Nepal Health Research council for providing raw data. The author is grateful to University of Cyberjaya for providing necessary assistance for the study.

## Authors Contribution

**Conceptualization:** Bhawana Bhandari, Suresh Mehata, Datin Mariyani Ahamad Niruzzudin.

**Formal Analysis:** Bihumgum Bista, Bhawana Bhandari

**Funding Support:** University Grant Commission

**Funding aquistition:** Bhawana Bhandari

**Investigation:** WHO STEPS Survey 2019

**Project administration:** Megnath Dhimal, Bihumgum Bista

**Methodology:** Bhawana Bhandari, Suresh Mehata, Bihumgum Bista

**Software:** Bihumgum Bista

**Supervision:** Suresh Mehata, Datin Mariyani Niruzzadin Ahmad.

**Writing : Original draft:** Bhawana Bhandari

